# Evolution of SARS-CoV-2 Shedding in Exhaled Breath Aerosols

**DOI:** 10.1101/2022.07.27.22278121

**Authors:** Jianyu Lai, Kristen K. Coleman, S.-H. Sheldon Tai, Jennifer German, Filbert Hong, Barbara Albert, Yi Esparza, Aditya K. Srikakulapu, Maria Schanz, Isabel Sierra Maldonado, Molly Oertel, Naja Fadul, T. Louie Gold, Stuart Weston, Kristin Mullins, Kathleen M. McPhaul, Matthew Frieman, Donald K. Milton

## Abstract

Aerosol inhalation is increasingly well recognized as a major if not primary mode of transmission of SARS-CoV-2^1,2^. Over the course of the COVID-19 pandemic, three highly transmissible lineages evolved and became globally dominant^3^. One hypothesis to explain increased transmissibility is that natural selection favours variants with higher rates of viral aerosol shedding. However, the extent of aerosol shedding of successive SARS-CoV-2 variants is unknown. Here, we demonstrate that viral shedding (measured as RNA copies) into exhaled breath aerosol was significantly greater during infections with Alpha, Delta, and Omicron than with ancestral strains and variants not associated with increased transmissibility. The three highly transmissible variants independently evolved a high viral aerosol shedding phenotype, demonstrating convergent evolution. We did not observe statistically significant differences in rates of shedding between Alpha, Delta, and Omicron infections. The highest shedder in our study, however, had an Omicron infection and shed three orders of magnitude more viral RNA copies than the maximum observed for Delta and Alpha^4^. Our results also show that fully vaccinated and boosted individuals, when infected, can shed infectious SARS-CoV-2 via exhaled breath aerosols. These findings provide additional evidence that inhalation of infectious aerosols is the dominant mode of transmission and emphasize the importance of ventilation, filtration, and air disinfection to mitigate the pandemic and protect vulnerable populations. We anticipate that monitoring aerosol shedding from new SARS-CoV-2 variants and emerging pathogens will be an important component of future threat assessments and will help guide interventions to prevent transmission via inhalation exposure.

## Background

The transmissibility of severe acute respiratory syndrome coronavirus 2 (SARS-CoV-2) continues to increase as new variants emerge^5–7^. Three variants of concern (VOCs), Alpha (B.1.1.7), Delta (B.1.617.2), and Omicron (B.1.1.529), successively became dominant during 2021^3^. Each was identified as having increased transmissibility relative to earlier variants or ancestral strains^5–7^. The Alpha variant, first identified in the United Kingdom in September 2020, was designated a VOC by the World Health Organization on December 18, 2020^3^. The Delta variant was first documented in India in October 2020 and was listed as a VOC on May 11, 2021^3^. The Omicron variant, first identified in southern Africa in November 2021^8^, was designated a VOC on November 26, 2021^3^. Subvariants of Omicron remain the predominant variants worldwide.

The spread of SARS-CoV-2 VOCs appears to be driven by two independent attributes, enhanced transmissibility and partial immune escape^9^. For example, Omicron BA.1 is approximately three times more transmissible than Delta^10,11^, which may be driven by its ability to transmit among vaccinated and boosted populations with a relatively modest increase in transmissibility among unvaccinated persons^10^. However, the Omicron BA.2 subvariant is estimated to be 30-40% more transmissible than Omicron BA.1^12^, although it does not display enhanced antibody escape relative to BA.1^13,14^. Hui and colleagues recently demonstrated that Omicron BA.2 replicates more efficiently than BA.1 in human nasal and bronchial tissues^15^, which could partly explain the observed dominance of BA.2 over BA.1. Whether this change in replication competence impacts onward transmission via viral aerosol shedding is unknown.

Viral load in the upper respiratory tract, especially when characterized by culture rather than RNA levels, is generally thought to reflect infectiousness. Puhach *et al* ^16^ reported that, in unvaccinated individuals, the infectious viral load in upper respiratory samples was higher when infected with the Delta variant compared to other strains of SARS-CoV-2 prior to the evolution of VOCs. Surprisingly, however, in fully vaccinated individuals, the infectious viral load was lower when infected with Omicron than when infected with Delta. Given that Omicron outcompeted and replaced Delta, this suggests that something other than infectious viral load in the upper respiratory tract is driving the increased transmissibility of Omicron.

Multiple lines of evidence point to a central role for aerosol inhalation (i.e., airborne transmission) as the primary mode of SARS-CoV-2 transmission^1^. This suggests that VOCs associated with increased transmissibility may have been selected based on increased fitness for transmission via inhalation exposure. Aerobiological characteristics on which natural selection could operate to produce more transmissible variants include increased rate or duration of viral aerosol shedding and prolonged survival in the aerosolized state^17^. We previously reported that individuals infected with the Alpha variant shed viral RNA copies into *fine* aerosols (≤5 µm in diameter) at an 18-fold greater rate than did individuals infected with ancestral strains and variants not associated with increased transmissibility^4^. It is unknown whether the continued evolution of subsequent variants and subvariants that appear to be successively more transmissible is associated with continued increases in the rate of aerosol shedding. In this study, we report measurements of the rate of SARS-CoV-2 shedding into exhaled breath aerosol (EBA) by individuals with Delta and Omicron infections, compare the rates of viral aerosol shedding among Omicron subvariants, and describe the evolving rate of SARS-CoV-2 aerosol shedding over the course of the first two years of the pandemic using data from this and our previous publication on viral loads in EBA.

## Methods

COVID-19 cases and their close contacts from the University of Maryland and the surrounding community were recruited as part of an ongoing research project “StopCOVID@UMD” aiming to study the transmission of SARS-CoV-2^4^. This study was approved by the University of Maryland Institutional Review Board and the Human Research Protection Office of the Department of the Navy. Electronic informed consent was obtained and questionnaire data were collected and stored using REDCap^18^.

### Study population and data collection

Individuals with an active SARS-CoV-2 infection confirmed by a recent PCR test were recruited from June 6, 2020 to March 11, 2022. Data for participants enrolled from June 6, 2020 to April 30, 2021 were reported elsewhere^4^ and included for comparisons in our current data analyses. Basic demographic data, including vaccination information, were obtained from a baseline questionnaire. Participants were sampled one to thirteen days post-onset of symptoms. On each day of sample collection, the participants completed an online questionnaire to update the status of their symptoms and record any medication used. As previously described^4^, participants were asked to self-report 16 symptoms on a scale from zero to three. Separate composite symptom scores were then calculated for systemic, gastrointestinal, lower respiratory, and upper respiratory symptoms.

### Sample collection

During viral shedding assessment visits, participants provided saliva, mid-turbinate swabs (MTS), phone swabs, venous blood samples, and exhaled breath aerosol (EBA) samples collected with a Gesundheit-II (G-II) human exhaled bioaerosol collector^19^ following a loud speaking and singing protocol, as previously described^4^. Some participants completed two shedding assessment visits one to three days apart.

### Sample processing and laboratory analyses

Saliva was processed using the SalivaDirect method^20^. Nucleic acids were extracted from all other samples following the specific manufacturers’ protocols using the MagMax Pathogen RNA/DNA Kit (Applied Biosystems) on a KingFisher Duo Prime (Thermo Scientific). Viral RNA was detected and quantified using the TaqPath COVID-19 Multiplex Real-Time RT-PCR Assay. RNA copy numbers were reported per mL for saliva and per sample for all other sample types. The limit of detection (LOD, 95% probability of detection) was 62 copies/mL for saliva and 75 copies/sample for all other sample types. Aliquots of samples were sent to the University of Maryland School of Medicine for virus culture using TMPRSS2-expressing VeroE6 cells and A549-ACE2 cells as described in detail in our previous publication^4^. Plasma samples were assayed for antibodies to SARS-CoV-2 as previously described^4^. IgG antibodies were titered using the SARS-CoV-2 receptor binding domain (RBD) and nucleocapsid (N) proteins (ACRO Biosystems) as the targets. Genome sequencing of MTS samples was performed using a MinION sequencing system (Oxford Nanopore Technologies, ONT) following the 1200-bp tiled amplicon (“Midnight”) protocol^21^. Fastq reads were uploaded to the EPI2ME platform (ONT) for sequence assembly as well as clade and lineage analyses.

### Statistical analyses

Data cleaning and analyses were completed using R version 4.2.0 and RStudio^22^. Descriptive analyses were done for all participants and by time period of enrollment (from June 6, 2020 to April 30, 2021, and from September 14, 2021 to March 11, 2022). Boosted participants were defined as having received one vaccine booster dose no less than 8 days prior to study enrollment^23^.

The Mann–Whitney *U* Test was used for pairwise comparisons of EBA viral RNA loads and number of coughs per 30-minute session for Alpha, Delta, Omicron, and other variants, for pairwise comparisons of composite symptom scores and individual symptoms for Alpha, Delta, Omicron BA.1, Omicron BA.2, and other variants, and to compare EBA viral RNA load by booster and MTS viral RNA load by anti-nucleocapsid IgG status for Omicron cases. The Kruskal-Wallis test was used to compare EBA viral RNA loads among three subvariants of Omicron (BA.1, BA.1.1, and BA.2) and for global comparison among variants in terms of EBA viral RNA load, number of coughs per 30-minute session, composite symptom scores, and individual symptoms. Spearman correlation coefficients (rho) and locally weighted smoothing (LOESS) curve with a 95% confidence interval were used to depict the correlation between EBA and MTS as well as EBA and saliva in terms of viral RNA copy numbers.

Linear mixed-effect models with censored responses^24,25^ (R package “lmec”^26^) were used to calculate the geometric means and standard deviations of viral RNA copy numbers for all sample types, and to estimate the effect of predictors on *fine* EBA viral load, accounting for censored observations below the limit of detection and nested random effects of subjects and repeated samples within subjects. Linear mixed-effect models (R package “lme4”^27^) were used for the effect of predictors on *coarse* EBA viral load because the models accounting for censored responses were unstable due to a large proportion of censored responses in some strata; a value of 1 was assigned to those with a viral load that was censored below the limit of detection (75 copies) in this analysis. All the adjusted models were selected based on the Akaike information criterion (AIC) while keeping age and sex for models over the course of the pandemic and keeping Omicron subvariants, age, and sex for models among participants with Omicron infections. Interactions between booster status and age, sex, as well as Omicron subvariants were considered in the adjusted models among the Omicron infections; only those that were included in the final models were presented.

## Results

From June 6, 2020 through March 11, 2022, we measured viral load in the exhaled breath of 93 individuals, 39% female and 61% male with mean age 25 years (range: 6 to 66 years; Extended Data Table 1), with a SARS-CoV-2 infection. Participants were mildly symptomatic (97%) or asymptomatic (3%) at the time of sampling. Persons enrolled from September 2021 through March 2022 had an active Delta or Omicron infection, were fully vaccinated, and had detectable IgG against the SARS-CoV-2 spike protein receptor binding domain. Among them, 20 (63%) were boosted prior to study enrolment, and 5 (16%) had detectable IgG against the SARS-CoV-2 nucleocapsid (N) protein (Extended Data Tables 1 and 2). Participants enrolled from June 2020 to April 2021 were previously reported^4^ and represent infections with ancestral strains, Alpha, and other variants (Gamma, Iota, and undetermined) prior to widespread vaccination.

Among the 3 Delta and 29 Omicron cases, we detected SARS-CoV-2 RNA in all sample types and recovered infectious virus from all sample types except fomites (Figure 1; Extended Data Table 3). A majority (21/32; 66%) of the participants with Delta and Omicron infections shed detectable concentrations of viral RNA in exhaled breath aerosol (EBA). Viral RNA loads in the *coarse* (>5 µm) and *fine* (≤5 µm) aerosol size fractions ranged from non-detect to 1.8×10^5^ and non-detect to 1.8×10^7^ RNA copies per 30-minute EBA sample, respectively. The viral RNA load in the *fine* fraction was on average five times greater than in the *coarse* fraction and accounted for most of the total exhaled viral RNA load.

**Figure 1.**
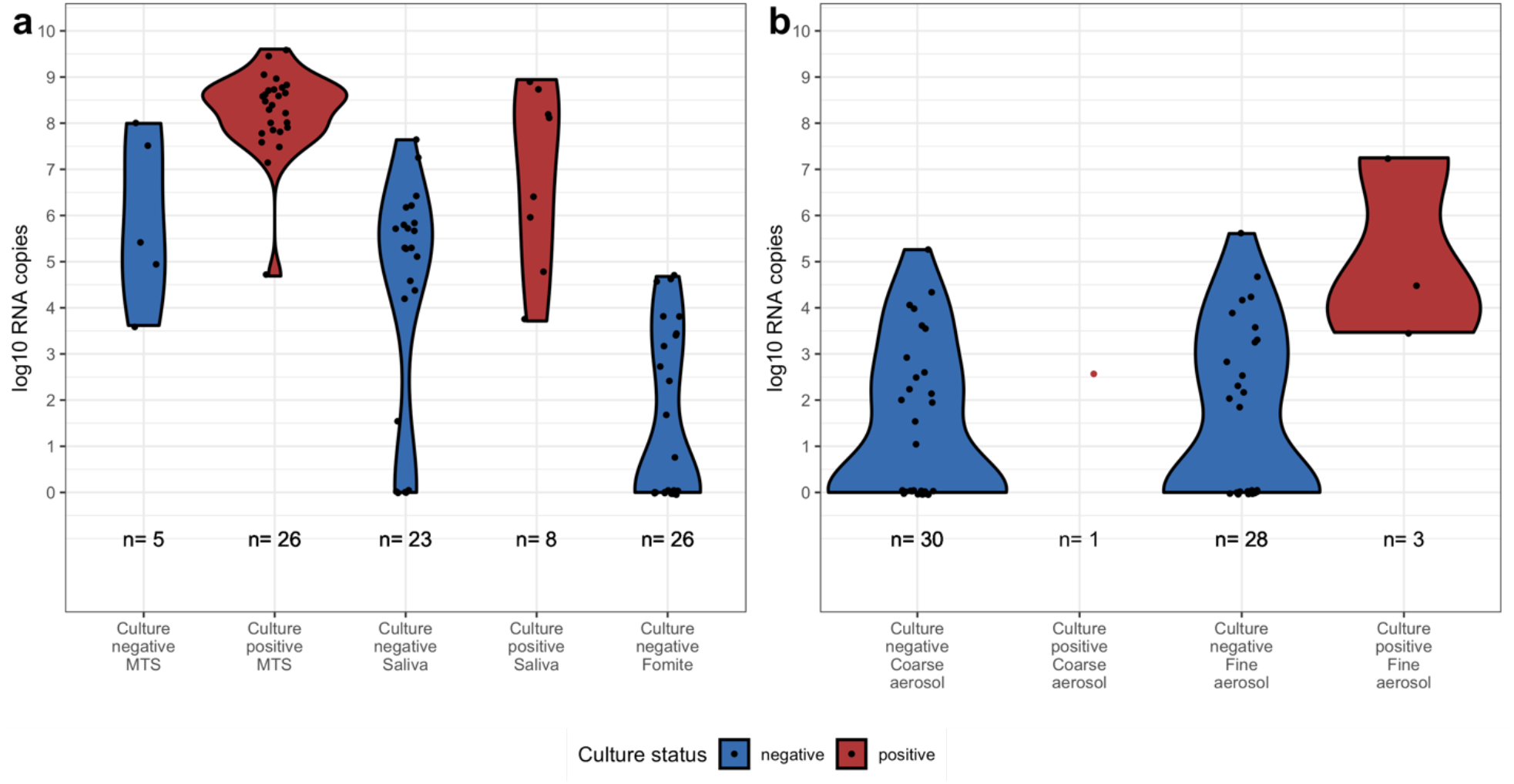
Viral RNA load and culture results from SARS-CoV-2 Delta and Omicron infections. Violin plots present the viral RNA copies on the log 10 scale of cultured negative and positive samples from SARS-CoV-2 Delta and Omicron infections by sample type from September 14, 2021 to March 11, 2022. Each point represents a sample. **a**, Mid-turbinate swab (MTS), saliva, and fomite samples. Fomite means swab of participant’s mobile phone. **b**, *Coarse* (>5 µm in diameter) and *Fine* (≤5 µm in diameter) exhaled breath aerosol (EBA) from 30-minute sampling events. The *n* at the bottom of the plots indicates the number of samples. Samples with no detectable viral RNA were assigned a copy number value of one.

### SARS-CoV-2 aerosol shedding during Delta variant infections

We detected viral RNA in EBA from two (66.7%) of the participants with Delta infections, each yielding positive virus cultures from EBA. From one of these individuals, fully vaccinated with NVX-CoV2373, we cultured SARS-CoV-2 from an EBA *fine* aerosol fraction that contained 3.0×10^4^ RNA copies. For the other individual, fully vaccinated with BNT162b2, we cultured virus from an EBA *coarse* aerosol fraction that contained 3.6×10^2^ RNA copies. None of the participants with Delta infections were boosted at the time of infection.

### SARS-CoV-2 aerosol shedding during Omicron (BA.1, BA.1.1, and BA.2) infections

Among participants with Omicron infections, 19/29 (66%) shed detectable concentrations of viral RNA into EBA. Two Omicron infected participants (both with BA.1.1) yielded positive virus cultures from their EBA *fine* aerosol size fraction. One was fully vaccinated (not boosted) with BNT162b2 and emitted the highest level of viral RNA in a *fine* EBA sample (1.8×10^7^ copies) among all 93 individuals studied over the course of the pandemic. The other individual, fully vaccinated with BNT162b2 and boosted with BNT162b2 more than two weeks prior to infection, shed 2.9×10^3^ viral RNA copies into their EBA *fine* aerosol fraction.

*Fine* EBA viral RNA loads from Omicron infections were, on average, similar to those from Alpha and Delta infections (Figure 2). However, the maximum Omicron *fine* EBA shedding rates tended to be higher while maximum mid-turbinate swabs (MTS) viral RNA loads tended to be lower compared to earlier highly transmissible VOCs (Figure 3). Omicron MTS viral RNA load was a weak positive correlate of *fine* EBA viral RNA load (rho = 0.36, p = 0.015), in contrast to ancestral strains and other variants where MTS load was moderately positively correlated with EBA load (rho = 0.59, p < 0.0001; Figure 3). Omicron viral RNA loads in saliva, however, trended toward a stronger, albeit still moderate, correlation with EBA load (rho = 0.58, p < 0.0001) when compared with earlier strains and variants (rho = 0.41, p < 0.0001). A similar pattern was observed for *coarse* aerosol viral RNA (Extended Data Figure 1).

**Figure 2.**
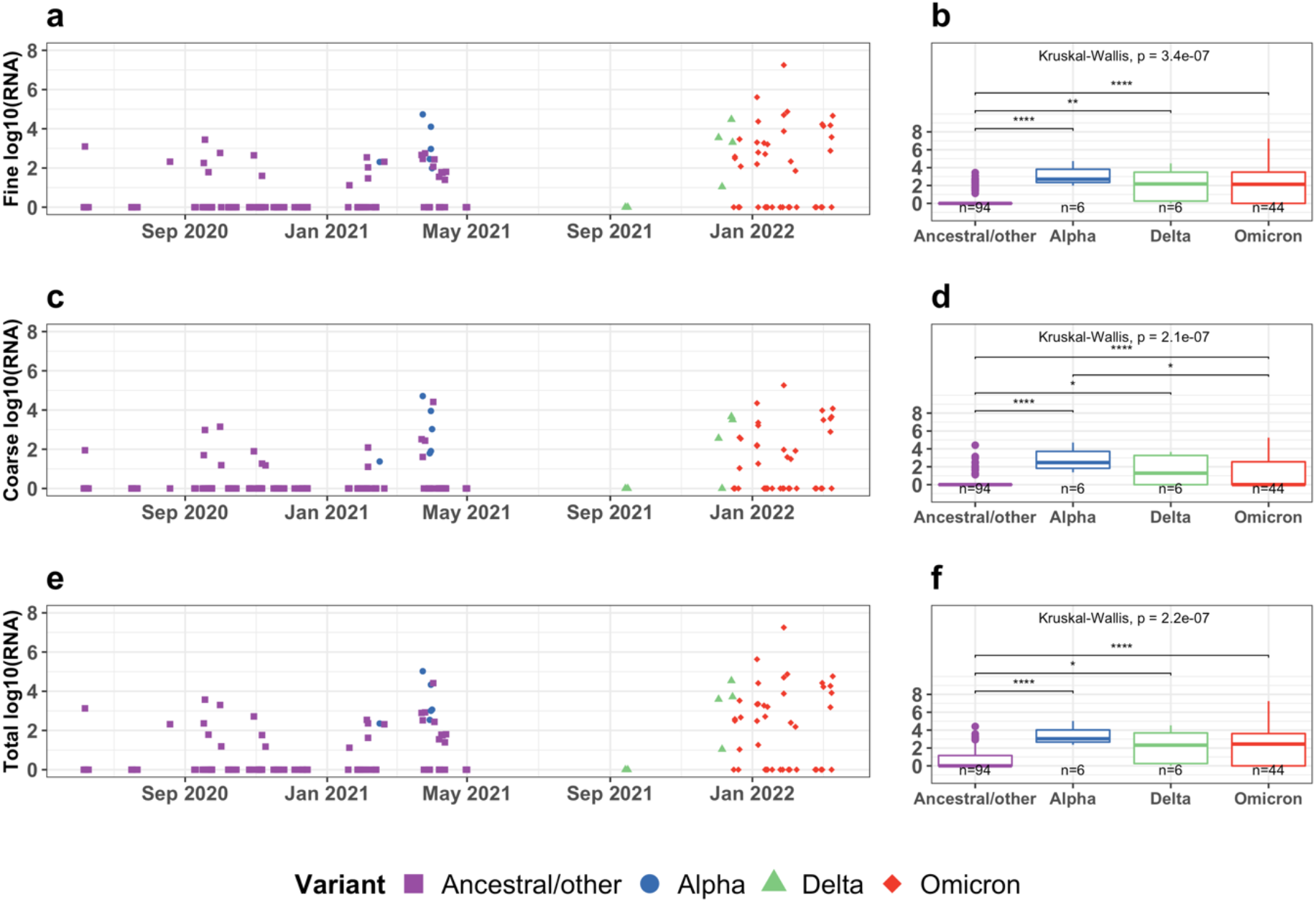
Viral RNA copies (log 10 scale) in exhaled breath aerosol (EBA) samples for SARS-CoV-2 variants over time. **a, c, e**, Scatter plots depict the change of viral RNA copies on the log 10 scale from June 6, 2020 to March 11, 2022. Each point represents a sample collected for an individual on a specific date. **b, d, f**, Boxplots present the comparison of viral RNA copies on the log 10 scale by SARS-CoV-2 variants. The Kruskal-Wallis p-value indicates the global comparison among the four variants. The asterisks indicates the pairwise comparison between two variants. Only those with a p-value less than 0.05 are shown (*: p <= 0.05; **: p <= 0.01; ***: p <= 0.001; ****: p <= 0.0001). The *n* indicates the number of samples included in each boxplot. **a, b**, Fine EBA (≤5 µm in diameter); **c, d**, Coarse EBA (>5 µm in diameter); **e, f**, Total EBA (fine and coarse combined). *Ancestral/other* means SARS-CoV-2 ancestral strains and other variants not associated with increased transmissibility.

**Figure 3.**
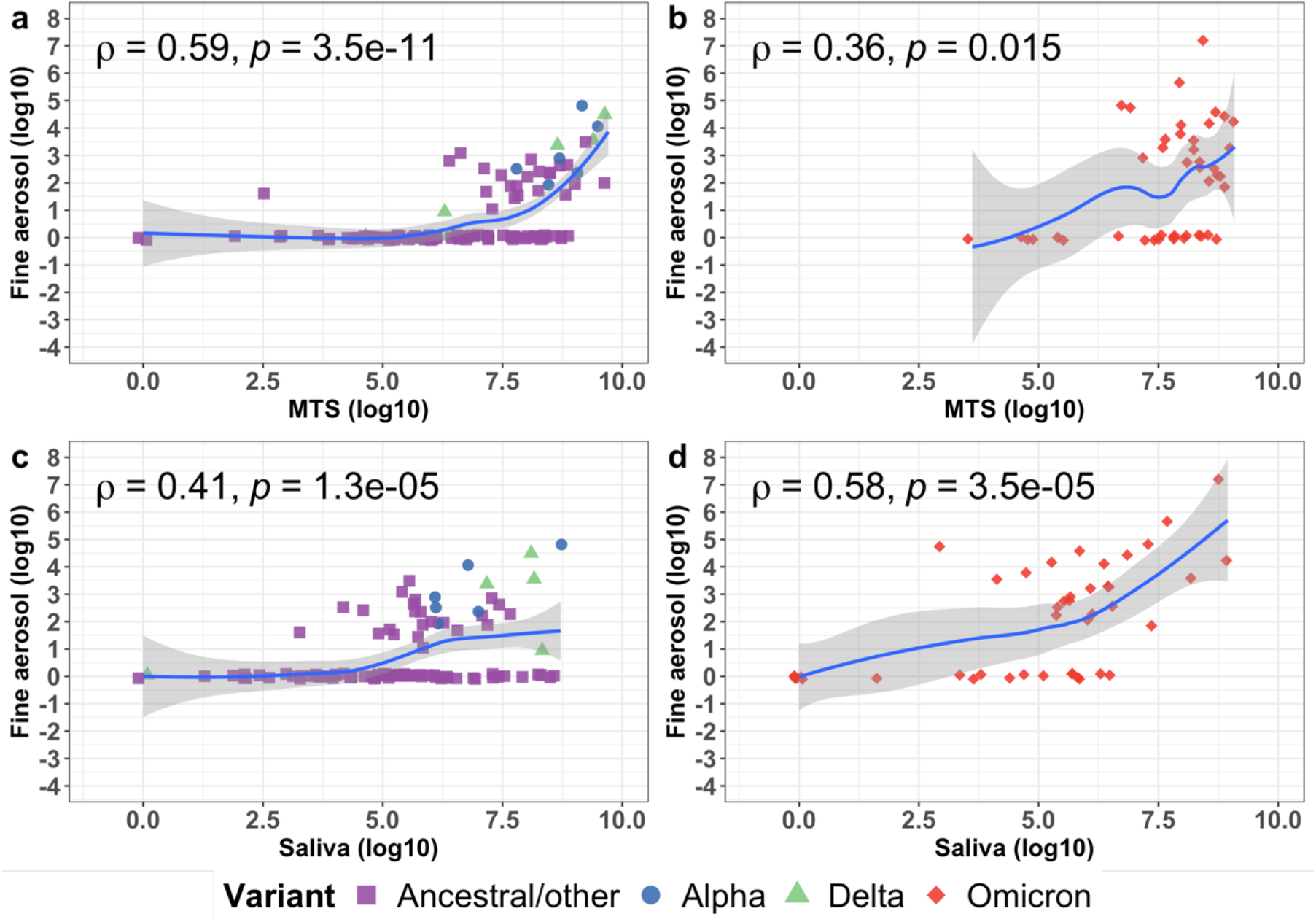
Correlation between viral RNA copies in fine (≤5 µm in diameter) exhaled breath aerosol (EBA) and mid-turbinate swab (MTS) samples as well as saliva. The locally weighted smoothing (LOESS) curves demonstrate the correlation of the RNA copies on the log 10 scale between *fine* EBA and MTS (**a** and **b**) as well as *fine* EBA and saliva (**c** and **d**) from June 6, 2020 to March 11, 2022. The shaded areas represent the 95% confidence interval of the smooth curves. Each point represents samples collected from an individual on a specific day. Rho (ρ) means spearman correlation coefficient. **a** and **c** depict the correlations among Pre-Omicron (ancestral/other, Alpha, and Delta) infections. **b** and **d** depict the correlations among Omicron (including BA.1, BA.1.1, BA.2) infections. *Ancestral/other* means SARS-CoV-2 ancestral strains and other variants not associated with increased transmissibility.

We did not observe a significant difference in viral aerosol shedding between Omicron BA.1, BA.1.1, and BA.2 (p>0.05; Extended Data Figure 2). Having received a vaccine booster dose was not associated with *fine* EBA viral RNA load (p = 0.97; Extended Data Figure 3 a-b). However, boosted individuals in our study shed higher loads of viral RNA in *coarse* EBA compared to non-boosted individuals (p=0.0056; Extended Data Figure 3 c-d). This effect was not evident for total EBA load (p = 0.81; Extended Data Figure 3 e-f), which was dominated by the *fine* aerosol fraction, as noted above.

Five participants with Omicron infections (one with BA.1, one with BA.1.1, and three with BA.2) were positive for anti-nucleocapsid (anti-N) protein IgG in sera at the time of enrolment, one to six days post symptom onset. Four of the five had received a booster >8 days prior to onset of symptoms. Two reported prior infection(s); two denied prior infection and one did not respond to questions about prior infection. We detected viral RNA in MTS samples from all five. Their MTS, however, contained significantly lower RNA copy numbers than Omicron infections in the absence of anti-N IgG (p=0.00045; Extended Data Figure 4). These five are the only Omicron infections that yielded culture negative MTS samples (Figure 1). We detected viral RNA in saliva from only one of the five, the non-boosted case, and that sample was not culturable. None of the five shed detectable levels of SARS-CoV-2 RNA in EBA (limit of detection = 75 copies).

Three participants with Omicron infections (one BA.1, two BA.1.1) were children aged 6-12 years. One *coarse* EBA sample contained a trace amount of SARS-CoV-2 RNA. None of the children’s *fine* EBA samples contained detectable levels of RNA. MTS samples from each of the three children yielded positive virus cultures. All of their saliva and EBA samples were culture negative.

### Predictors of viral aerosol shedding from Omicron infections

Among the 29 Omicron cases, higher saliva viral RNA load, systemic symptom score, and number of coughs per 30-minute sampling session were significant predictors for higher *fine* EBA viral RNA load in a model adjusted for age, sex, and subvariant BA.2 compared with BA.1 and BA.1.1 (Figure 4 a-b). Only higher saliva viral RNA load and systemic symptom score were significant predictors for higher *coarse* EBA viral RNA load in an adjusted model (Extended Data Figure 5 a-b). The BA.2 subvariant was not associated with significantly greater shedding into either *fine* or *coarse* EBA compared to BA.1 and BA.1.1.

**Figure 4.**
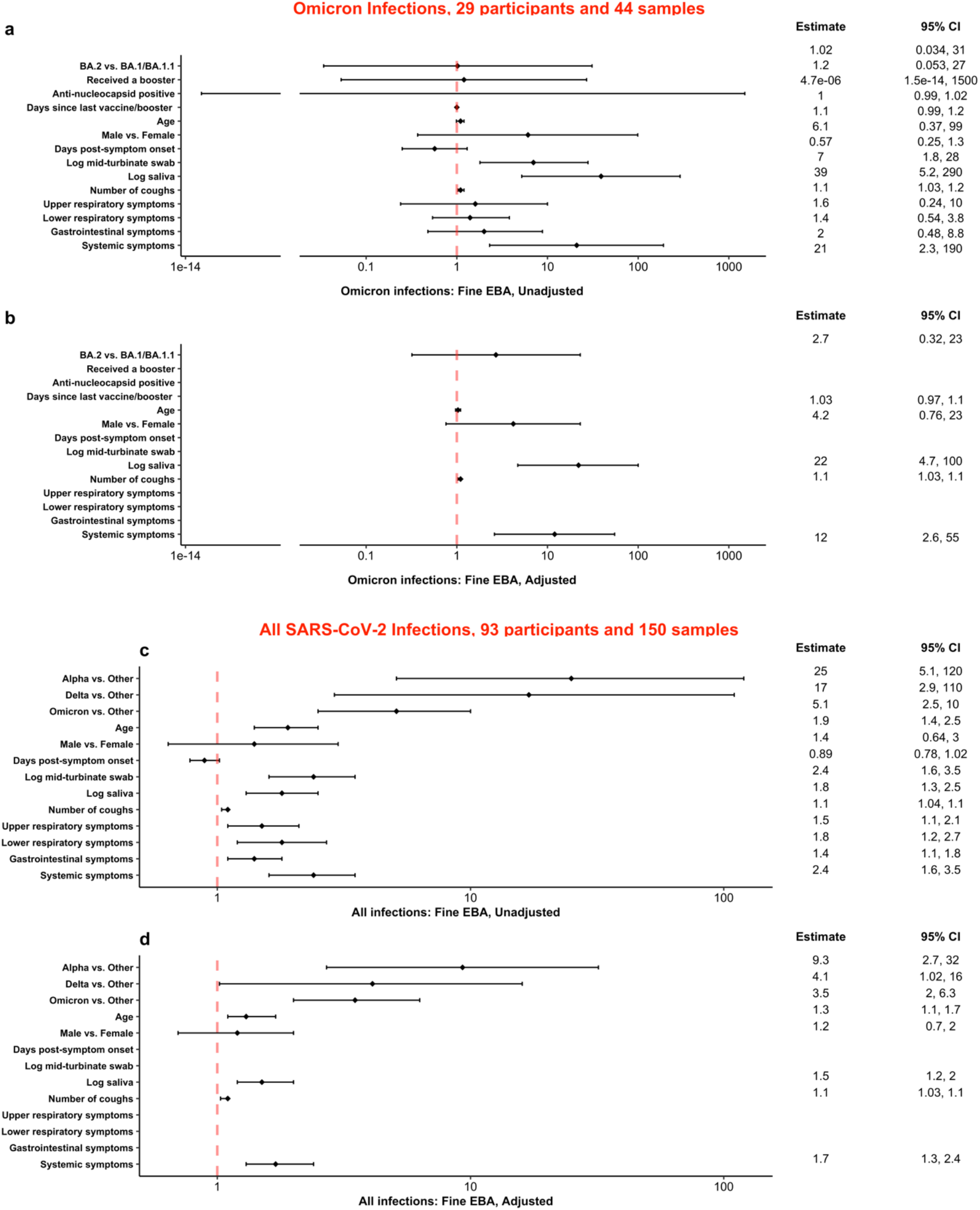
Predictors for SARS-CoV-2 RNA loads in *fine* exhaled breath aerosol. **a-b**, Predictors for viral RNA loads in *fine* exhaled breath aerosol among 29 participants with Omicron infections enrolled from December 16, 2021 to March 11, 2022. **c-d**, Predictors of viral RNA loads in *fine* exhaled breath aerosol over the course of the pandemic from June 6, 2020 to March 11, 2022. Effect estimates and their 95% confidence intervals from linear mixed effect models accounting for censored responses below the limit of detection are shown as the ratio of RNA copy number of samples: variant to variants other than Alpha/Delta/Omicron, Omicron BA.2 to Omicron BA.1 and BA.1.1, received to not received a booster, anti-nucleocapsid positive to negative, male to female, or as the fold-increase in RNA copy number for a 10-year increase in age, 1-day increase in day post-symptom onset or days since last vaccine/booster, 1-count increase in numbers of coughs, and an interquartile range change in symptom scores, mid-turbinate swab and saliva RNA copy number.

### Evolution of SARS-CoV-2 aerosol shedding

Over the course of the pandemic (Figure 4 c-d), three highly transmissible SARS-CoV-2 variants (Alpha, Delta, or Omicron), as well as higher systemic symptom score, saliva viral RNA load, age, and number of coughs per 30-minute sampling session were significant predictors for higher *fine* EBA viral RNA load in an age and sex adjusted model. Highly transmissible VOCs were associated with increased *coarse* aerosol shedding in unadjusted analyses but were not significant predictors in adjusted models. Higher systemic symptom score, MTS viral RNA load, and age were significant predictors for higher *coarse* EBA viral RNA load in an adjusted model controlling for age and sex (Extended Data Figure 5 c-d).

Participants with Delta and Omicron infections on average coughed more per 30-minute sampling session compared to Alpha, ancestral strains, and other variants (Extended Data Figure 6, a-b). The highest cough count was from a BA.1.1 case who coughed 69 times during the 30-minute sampling session. Two participants (one infected with Omicron BA.2 and one with ancestral strain, B.1.509) sneezed during the sampling sessions, each sneezing once. Omicron BA.1 (including BA.1.1) and BA.2 cases generally reported more upper and lower respiratory symptoms compared to those infected with ancestral strains and other variants (Extended Data Figure 6-7).

## Discussion

Our observations demonstrate that fully vaccinated persons infected with SARS-CoV-2 Delta, and vaccinated and boosted persons infected with Omicron variants, shed infectious viral aerosols. We show that infection caused by Alpha, Delta, and Omicron (i.e., variants associated with increased transmissibility) produced significantly greater viral aerosol shedding (measured as RNA copies) than did infections with ancestral strains and variants not associated with increased transmissibility. These data indicate that a characteristic of highly transmissible variants is a high rate of viral shedding into aerosols. These three highly transmissible variants represent three distinct SARS-CoV-2 clades that independently evolved high shedding phenotypes. This evidence for convergent evolution of increased viral aerosol shedding is consistent with a dominant role for airborne transmission (defined as inhalation of viral aerosols regardless of distance that the aerosol traversed) in the spread of COVID-19^1^.

The highest viral EBA shedder in our study was a person with an Omicron infection whose *fine* EBA sample contained 1.8×10^7^ RNA copies, a value three orders of magnitude higher than the maximum values observed for Delta and previously reported Alpha variant infections,^4^ and 2.4-fold less than the maximum value we previously observed for influenza^28^. Among the three highly transmissible variants, however, we did not observe statistically significant differences in the geometric mean rates of viral RNA shedding into EBA. This may suggest that variants prone to result in more extreme supershedding outlier infections, a property not well-characterized by the geometric mean, could exhibit increased transmissibility through superspreading. Thus, superspreading as a biological factor, not only as a result social behaviour^29^, may be a driving force behind dominance of new variants when the variants differ minimally regarding immune escape. The geometric mean RNA copy number in EBA was two orders of magnitude less than we observed for symptomatic influenza cases, suggesting that future SARS-CoV-2 variants associated with higher rates of viral aerosol shedding may yet arise.

The *fine* aerosol fraction (≤5 µm) consistently contained greater numbers of viral particles based on RNA copy number compared to the *coarse* aerosol fraction (>5 µm), and dominated the total aerosol load in all of the SARS-CoV-2 infections studied throughout the pandemic using a well-characterized breath aerosol collector^4,30,31^. This pattern mirrored results from earlier studies of influenza^28,32–35^. These observations are consistent with data showing that bubble film burst due to airway closure and reopening is the dominant mechanism of respiratory aerosol generation^36–38^ and that bubble films concentrate microorganisms relative to their concentration in bulk fluids by orders of magnitude^39–41^. When considered together with the relatively more efficient concentration and aerosolization of enveloped compared with naked protein capsid viruses,^42^ it is perhaps not surprising that respiratory viral pandemics of the last >100 years have been caused by enveloped viruses. Furthermore, selection may be favouring variants that replicate more efficiently at sites where aerosols are generated. Hui *et al* ^43^ found that Omicron variants replicated to 70-fold higher titres in human bronchial *ex vivo* cultures than wild-type or Delta strains at 24 and 48 hours after infection, indicating that Omicron infections result in higher viral loads in conducting airways including potentially small airways where reopening film burst aerosol generation occurs.

While mutations in the SARS-CoV-2 spike protein have allowed Omicron to partially evade infection- and vaccine-acquired immunity^44^, our findings together with Hui *et al* ^43^ suggest that high viral aerosol shedding is also contributing to Omicron’s high transmission rates. This is in direct alignment with a mathematical model by Bushman *et al* ^9^ demonstrating that only variants exhibiting immune evasive properties *and* enhanced transmissibility can thrive in populations as immunity from infection and vaccination increases.

Puhach *et al* ^16^ demonstrated that fully vaccinated Omicron cases had lower infectious viral loads in their nasopharynx compared to fully vaccinated Delta cases, and concluded that Omicron’s transmission advantage was likely not caused by higher infectious viral loads in the upper respiratory tract, as characterized by nasopharyngeal swabs (NPS). Because viral loads in aerosol samples tend to be low, we used culture methods designed to optimize sensitivity at the expense of quantification of infectious virus and cannot directly compare our culture results. However, our observations suggest that an explanation for this finding may be a poor correlation of viral load in the upper respiratory tract, as characterized by NPS or MTS, with viral aerosol shedding. We observed, in contrast to Puhach *et al* ^16^, that viral RNA aerosol shedding was similar for Delta and Omicron. The two highest viral aerosol shedders we studied had Omicron infections, despite having relatively low viral RNA loads in their MTS.

We previously reported that, for infections studied through April of 2021, high MTS viral RNA load was a strong risk factor for high viral RNA load for both *coarse* and *fine* EBA fractions^4^. With Omicron, however, we see a clear shift toward saliva being a stronger predictor of the viral RNA load in EBA. This was evident for both *coarse* and *fine* EBA viral RNA load in our linear mixed-effects models for Omicron infections and can be clearly seen in our correlation plots. These data suggest that viral RNA load in saliva might be a better predictor of contagiousness than MTS.

Omicron BA.2 appeared to be more transmissible than BA.1 in a study of Danish households^6^. However, the reported increase in transmissibility of BA.2 over BA.1 was limited to unvaccinated primary cases; fully vaccinated and boosted primary cases infected with BA.2 were significantly less likely to transmit BA.2 than BA.1^12^. Antibody escape is not thought to be responsible for the dominance of BA.2 over BA.1^13,14^. One recently observed advantage of BA.2 is an increased competence for replication in human nasal and bronchial tissues^15^. Thus far, this change does not appear to impact average viral aerosol shedding rates among vaccinated/boosted individuals with Omicron breakthrough infections as we did not see evidence of a significant difference in viral RNA aerosol shedding between people infected with BA.1, BA.1.1 and BA.2. Given that the dominance of BA.2 seems to have been associated with transmission by unvaccinated individuals, we might expect to see increased aerosol shedding from unvaccinated infected persons. Our data cannot address that possibility as all Omicron cases in our study were fully vaccinated and some were boosted. Recent data indicate newer Omicron variants, particularly BA.2.12.1, BA.4 and BA.5 appearing after the end of the current study, are even more transmissible and are able to escape antibody neutralization elicited by both vaccination and prior Omicron infection^45–47^. One possibility to account for the increased transmission in these new variants is that reduced antibody neutralization may contribute to higher airway viral loads and thus more viral aerosol shedding. Further investigation is needed to understand how infection with evolving antibody escape variants may impact aerosol shedding.

We observed that five participants with an Omicron infection were positive for anti-N protein IgG at the time of enrolment of whom two reported prior infections. Since the median time of seroconversion for anti-N IgG is 10 days^48^, anti-N IgG wanes with an estimated half-life of 85 days^49^, and the anamnestic response requires 3-4 days to first become detectable^50,51^, it is possible that the remaining three participants had experienced a prior asymptomatic infection with a rapid humoral response to their current infection or had been infected earlier than indicated by their reported symptom onset. The presence of anti-N IgG may indicate a broad immune response to infection, including IgA secretion. Infection produces a more robust IgA response than intramuscular vaccination^52^, and IgA concentrations decline more slowly after infection than those of IgG^53^. IgA is a potent neutralizer of SARS-CoV-2 during early infection^54^. These participants had no PCR-detectable levels of virus in EBA, on fomites and all but one saliva sample, and the viral RNA load in their MTS was significantly lower than that of the other Omicron cases. These observations together suggest that antibody responses involving both IgG and IgA in these participants may have played a role in reducing viral loads overall and limiting shedding in EBA. However, as recent successive Omicron subvariants are able to escape antibody neutralization elicited by prior Omicron infection,^45–47^ we might not expect to observe such a reduction in viral aerosol shedding among seropositive individuals infected with future variants.

*Fine* EBA viral RNA loads did not differ by vaccine booster status among Omicron cases, however, boosted individuals shed higher loads in *coarse* EBA compared to non-boosted individuals. The effect was observed only among cases who did not have evidence of recent infection based on anti-N titres. This unexpected finding is not likely to impact transmissibility because the total EBA load, dominated by the *fine* EBA load, was not associated with booster status. It raises questions, however, about possible selection bias and differential impact of prior immunity from vaccination and infection on viral aerosol shedding from different anatomical sites within the respiratory tract. *Coarse* aerosol in exhaled breath is generated in the upper respiratory tract^55^. The upper respiratory tract including large intrathoracic airways are secretory-IgA dominant environments whereas the terminal bronchioles, the site of *fine* aerosol generation by airway reopening, are an IgG dominant environment^56,57^. Sheikh-Mohamed et al., recently reported that approximately 30% of individuals produce and maintain stable levels of anti-Spike/RBD IgA in serum and saliva in response to SARS-CoV-2 mRNA vaccine^52^ and that higher post vaccination IgA levels are associated with protection against breakthrough infection^52^. Because all the Omicron cases we studied were vaccinated (i.e., breakthrough infections), this heterogeneity in IgA response to vaccination introduces a potential selection bias among our study’s boosted population, if getting boosted is less likely for people with strong IgA responses to vaccination or a more distant history of COVID-19 and higher specific IgA in their mucosa. Further studies measuring the potential effect of IgA levels on the intensity and size distribution of viral aerosol shedding are warranted.

Our study has several limitations. Although we recruited throughout the pandemic, our sample size for each variant and subvariant was relatively small. As a result, we were limited in making comprehensive comparisons such as the correlation between EBA viral RNA load and culture positivity for specific variants. Although we were able to sample children infected with the Omicron variant, our sample size was too small to make any conclusions about viral aerosol shedding from children. The EBA collection procedure is not suitable for children under age 6 years. Lastly, we did not sample participants throughout their entire infection. Because viral loads in aerosol samples were low, we opted for a sensitive but non-quantitative measure of infectiousness. Thus, we were unable to assess the impact of variants and Omicron subvariants on the duration of viral aerosol shedding and infectious virus titres in EBA.

In conclusion, our findings demonstrate that infected persons shed infectious SARS-CoV-2 aerosols even when fully vaccinated and boosted. Evolutionary selection appears to have favoured SARS-CoV-2 variants associated with higher viral aerosol shedding. Comparison with shedding rates for influenza suggests that continued evolution of still higher aerosol shedding rates may be possible. The combination of immune evasive properties *and* high viral aerosol shedding were likely responsible for Omicron’s rapid spread and replacement of the Delta variant, even as infection- and vaccine-acquired immunity increased. Thus, non-pharmaceutical interventions, especially indoor air hygiene (e.g., ventilation, filtration, and air disinfection with germicidal UV) and targeted masking and respirators, are still needed to mitigate COVID-19 transmission in vaccinated communities to prevent post-acute COVID-19 sequalae^58^ and to protect vulnerable populations.

## Data Availability

Deidentified data for the accepted manuscript will be made available on the Open Science Framework repository.

## Data and Code Availability

Deidentified data for the accepted manuscript will be made available on the Open Science Framework repository. Custom code used to analyse the data will be made available on a public github repository with linkage to the Open Science Framework repository.

## Author contributions

D.K.M. conceived the project and obtained funding.

D.K.M., F.H., B.A., Y.E., J.L., S.S.T., J.G., I.S.M. conceptualized the project and designed the study.

I.S.M., A.K.S., M.O., and N.F. recruited study volunteers.

K.K.C., A.K.S., and M.O. collected exhaled breath samples.

B.A., Y.E., J.L., K.M.M., M.O., and N.F. collected clinical samples.

S.S.T., J.G., and MS processed and analysed samples.

S.W. and M.F. performed virus culture.

K.M. performed antibody tests on sera samples.

F.H. performed data management and curation.

J.L. performed data analyses.

J.L., K.K.C., and T.L.G. drafted the original manuscript.

All authors participated in reviewing and editing the manuscript.

## Acknowledgements

We thank all the other members and previous staff of the University of Maryland StopCOVID Research Group for their efforts in recruiting participants and sample collection and processing: Oluwasanmi Oladapo Adenaiye, P. Jacob Bueno de Mesquita, Aaron Kassman, Michael Lutchenkov, Dewansh Rastogi, Delwin Suraj, Faith Touré, Rhonda Washington-Lewis, Somayeh Youssefi, Mara Cai, Ashok Agrawala. We also thank Dr. Tianzhou Ma from the Department of Epidemiology and Biostatistics, University of Maryland School of Public Health, College Park, for providing advice on statistical modeling, and Dr. Jamal Fadul and his clinic in College Park, Maryland, for assistance in recruiting study participants.

