## Extended Data Figures and Tables for "Evolution of SARS-CoV-2 Shedding in Exhaled Breath Aerosols"

**Extended Data Table 1.** Demographics for SARS-CoV-2 cases enrolled June 6, 2020 – March 11, 2022

| | | Enrolled September 2021 - March 2022 | Enrolled June 2020 - April 2021[a] | All participants |
|---|---|---|---|---|
| **Number of participants** | | 32 | 61 | 93 |
| **Number of exhaled breath samples** | | 50 | 100 | 150 |
| **Variant, N (%)** | **Ancestral strains and other** | 0 (0) | 57 (93) | 57 (61) |
| | **Alpha** | 0 (0) | 4 (7) | 4 (4) |
| | **Delta** | 3 (9) | 0 (0) | 3 (3) |
| | **Omicron BA.1** | 8 (25) | 0 (0) | 8 (9) |
| | **Omicron BA.1.1** | 14 (44) | 0 (0) | 14 (15) |
| | **Omicron BA.2** | 7 (22) | 0 (0) | 7 (8) |
| **Female, N (%)** | | 13 (41) | 23 (38) | 36 (39) |
| **Age, mean ± SD** | | 27.2 ± 15.3 | 23.6 ± 9 | 24.8 ± 11.6 |
| **Age group, N (%)** | **<18** | 3 (9) | 1 (2) | 4 (4) |
| | **18-45** | 24 (75) | 57 (93) | 81 (87) |
| | **>45** | 5 (16) | 3 (5) | 8 (9) |
| **Race/Ethnicity, N(%)** | **White** | 19 (59) | 48 (79) | 67 (72) |
| | **Black/African American** | 5 (16) | 7 (12) | 12 (13) |
| | **Hispanic** | 5 (16) | 8 (13) | 13 (14) |
| **BMI, mean ± SD** | | 24.7 ± 5.4 | 25.2 ± 4.5 | 25 ± 4.8 |
| **Chronic respiratory illness, N (%)[b]** | | 6 (19) | 13 (21) | 19 (20) |
| **Vaccination status, N (%)[c]** | **Boosted** | 20 (63) | 0 (0) | 20 (22) |
| | **Fully vaccinated, not boosted** | 12 (37) | 0 (0) | 12 (13) |
| | **Partially vaccinated** | 0 (0) | 3 (5) | 3 (3) |
| | **Not vaccinated** | 0 (0) | 58 (95) | 58 (62) |
| **Anti-spike RBD antibody (IgG), N (%)** | | 32 (100) | 6 (10) [d] | 38 (41) |
| **Anti-nucleocapsid antibody (IgG), N (%)** | | 5 (16) | N/A[e] | 5 (5) |
| **Ever symptomatic, N (%)** | | 32 (100) | 58 (95) | 90 (97) |
| **Symptomatic participants** | **Days post symptom onset[f], mean ± SD (range)** | 3 ± 2 (1-7) | 5 ± 3 (0-13) | 4 ± 2 (0-13) |
| | **Coughs per 30 min, mean ± SD (range)** | 8 ± 15 (0-69) | 1 ± 4 (0-24) | 4 ± 10 (0-69) |
| | **Median upper respiratory symptoms[g] (IQR)** | 3.5 (2 - 6) | 2 (1 - 3.8) | 3 (1 - 4) |
| | **Median lower respiratory symptoms (IQR)** | 1 (0.2 - 2) | 0 (0 - 1.8) | 1 (0 - 2) |
| | **Median systemic symptoms (IQR)** | 2 (1 - 6) | 1 (0 - 3) | 2 (0 - 4) |
| | **Median gastrointestinal symptoms (IQR)** | 1 (0 - 2) | 0 (0 - 1) | 0 (0 - 1) |
| | **Temperature (C), mean ± SD** | 37 ± 0.3 | 37.2 ± 0.3 | 37.1 ± 0.3 |
| | **Oxygen saturation (SpO2), mean ± SD** | 97.9 ± 0.8 | 97.8 ± 1 | 97.8 ± 1 |

BMI = Body mass index; RBD = Receptor Binding Domain; IgG = Immunoglobulin class G; IQR = Interquartile range

a. Includes previously reported cases[2]
b. Chronic respiratory illness = volunteers with any chronic obstructive pulmonary disease, asthma, other lung diseases
c. Boosted = received one vaccine booster dose ≥ 8 days prior to study enrollment; Fully vaccinated, not boosted = received only two doses of BNT162B2, mRNA-1273, or NVX-CoV2373, or one dose of Ad26.COV2 ≥ 14 days prior to study enrollment; Partially vaccinated = received only one dose of BNT162B2 or mRNA-1273
d. Serologic status data for four participants were missing due to a lack of blood samples
e. Anti-nucleocapsid antibodies were not measured for these participants
f. Days since symptom onset at the time of each sample collection visit
g. Symptoms at the time of each sample collection visit. Sixteen symptoms were rated from 0 to 3 with a maximum possible composite score of 15 for upper respiratory, 9 for lower respiratory, 12 for systemic symptoms and 12 for gastrointestinal symptoms

**Extended Data Table 2.** Vaccine and booster types received by Delta and Omicron cases, September 14, 2021 – March 11, 2022.

| | | **Booster type** | | | |
|---|---|---|---|---|---|
| | | BNT162b2 | mRNA-1273 | NVX-CoV2373 | Not boosted |
| **Vaccine Type** | BNT162b2 | 9 | 5 | 0 | 10 |
| | mRNA-1273 | 1 | 3 | 0 | 0 |
| | Ad26.COV2.S | 1 | 1 | 0 | 1 |
| | NVX-CoV2373 | 0 | 0 | 0 | 1 |

**Extended Data Table 3.** SARS-CoV-2 in respiratory and fomite samples from Delta and Omicron participants, September 14, 2021 – March 11, 2022

| Sample type | Participants with ≥1 PCR positive sample, n/N (%)[a] | Participants with ≥1 culture positive sample, n/N (%)[b] | PCR Positive Samples, n/N (%)[c] | RNA copies per sample | |
|---|---|---|---|---|---|
| | | | | GM (95% CI)[d] | Maximum[e] |
| Saliva | 27/32 (84) | 8/31 (26) | 42/50 (84) | $6.6 \times 10^4$ ($9.5 \times 10^3$, $4.6 \times 10^5$) | $8.8 \times 10^8$ |
| Mid-turbinate Swab | 32/32 (100) | 26/31 (84) | 50/50 (100) | $3.6 \times 10^7$ ($1.3 \times 10^7$, $9.9 \times 10^7$) | $4.0 \times 10^9$ |
| Fomite (mobile phone screen) | 14/30 (47) | 0/26 (0) | 20/47 (43) | 16 (2.1, 130) | $4.5 \times 10^5$ |
| Coarse EBA[f] | 16/32 (50) | 1/31 (3) | 23/50 (46) | 29 (7.1, 120) | $1.8 \times 10^5$ |
| Fine EBA | 20/32 (62) | 3/31 (10) | 28/50 (56) | 150 (35, 600) | $1.8 \times 10^7$ |
| Total EBA (fine + coarse) | 21/32 (66) | 4/31 (13) | 51/100 (51) | 200 (52, 790) | $1.8 \times 10^7$ |

a. Number (n) of participants (N) with at least one sample ≥ the limit of detection (LOD, 62 copies/mL for saliva and 75 copies/sample for other sample types)
b. Number (n) of participants with samples subjected to culture (N) with at least one sample giving a positive culture for SARS-CoV-2
c. Number (n) of samples (N) ≥ LOD, with at least 1 replicate with confirmed amplification after inspection and quality control
d. GM = geometric mean. The GMs were computed by controlling for random effects of subject and sample nested within subjects and for censoring by the limit of detection using a linear mixed-effects model for censored responses (R package "lmec"[24])
e. The largest quantity of RNA copies detected based on the mean of replicate qRT-PCR aliquots
f. EBA = Exhaled breath aerosol. Each EBA sample was collected using a Gesundheit-II machine for 30 minutes

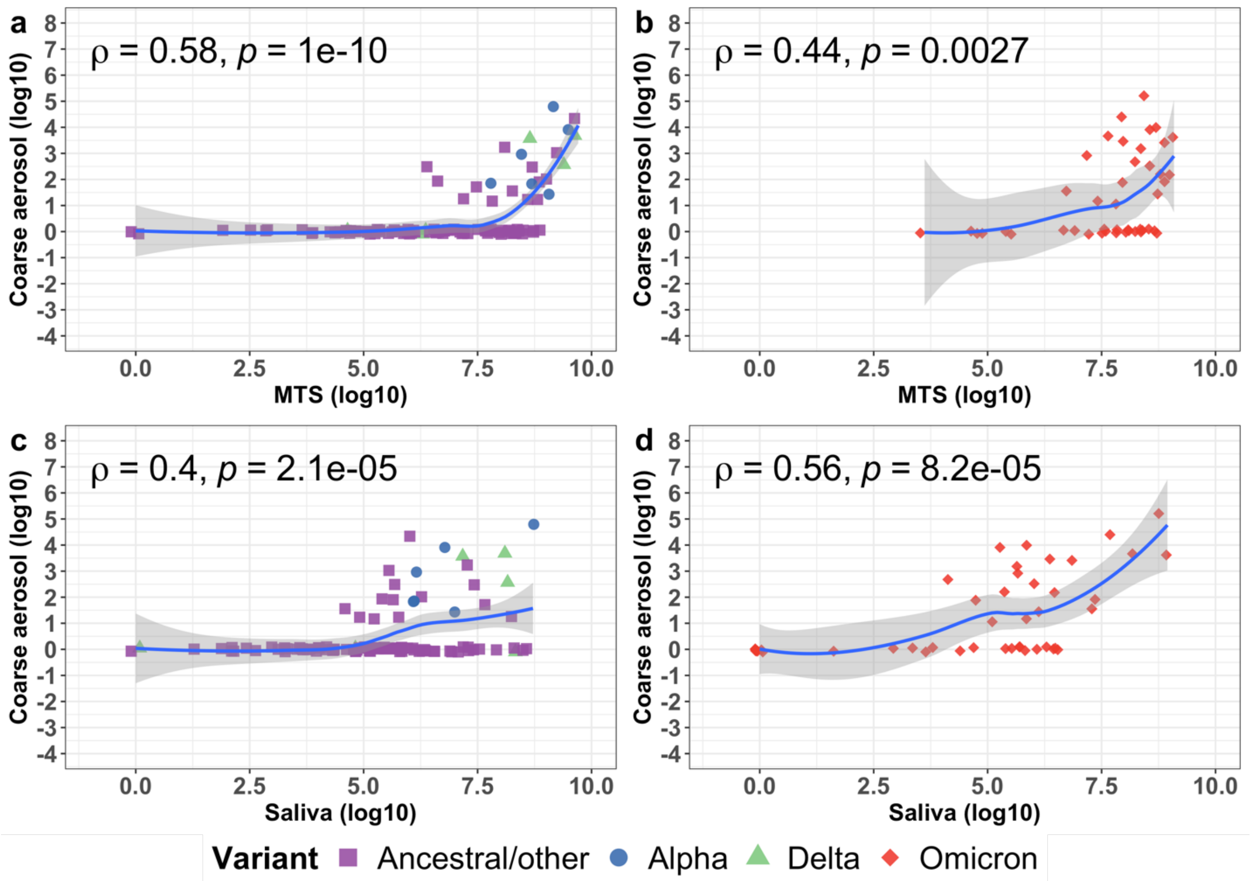


**Extended Data Figure 1. Correlation between SARS-CoV-2 RNA copies in coarse exhaled breath aerosol (>5 µm in diameter) and mid-turbinate swab (MTS) samples as well as saliva, June 6, 2020 – March 11, 2022.** The locally weighted smoothing (LOESS) curves demonstrate the correlation of the RNA copies on the log 10 scale between *coarse* EBA and MTS (**a**, **b**) as well as *coarse* EBA and saliva (**c**, **d**). The shaded areas represent the 95% confidence interval of the smooth curves. Each point represents samples collected from an individual on a specific day. Rho (ρ) is the Spearman correlation coefficient. **a** and **c** depict the correlations among pre-Omicron (ancestral/other, Alpha, and Delta) infections. **b** and **d** depict the correlations among Omicron (including BA.1, BA.1.1, BA.2) infections. *Ancestral/other* means SARS-CoV-2 ancestral strains and other variants not associated with increased transmissibility.

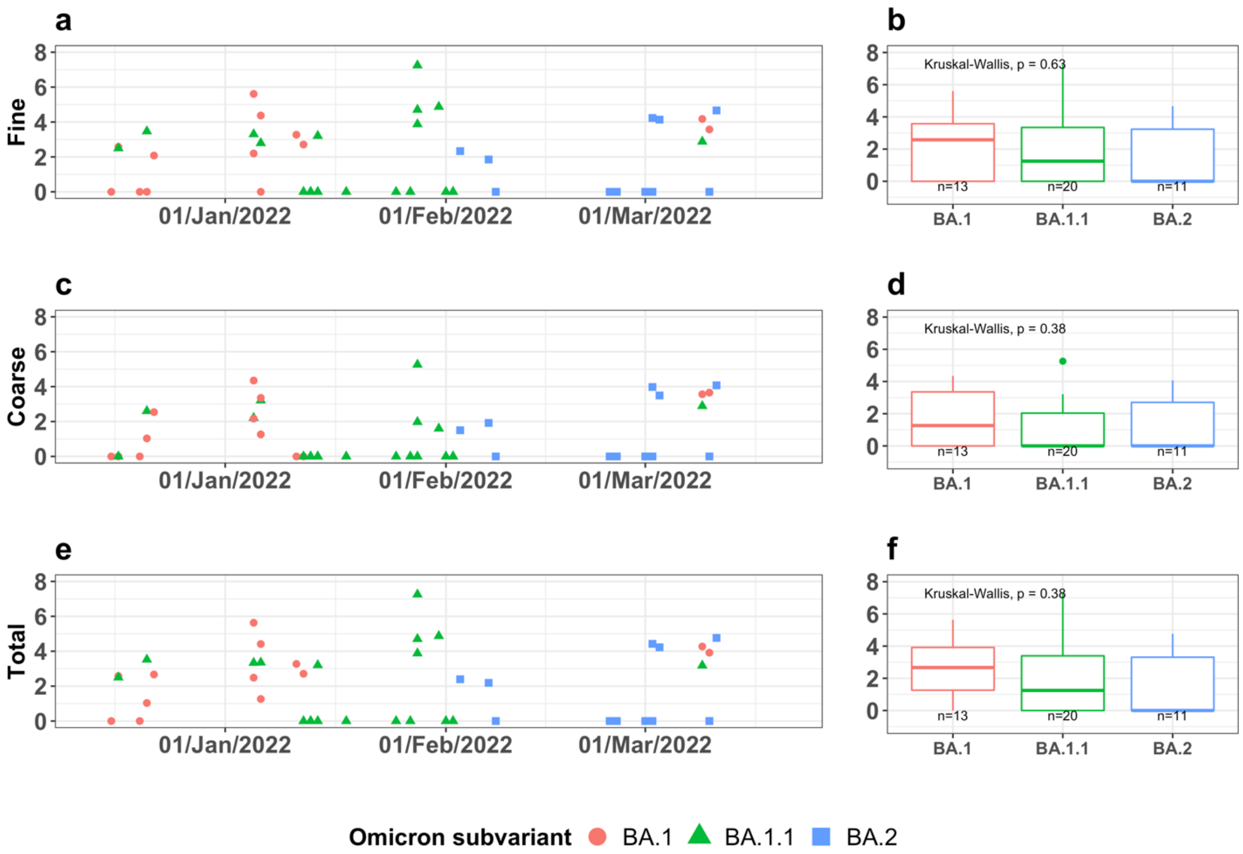


**Extended Data Figure 2. Viral RNA copies (log 10 scale) in fine and coarse exhaled breath aerosol (EBA) samples for SARS-CoV-2 Omicron subvariants over time, December 2021 – March 2022.**

**a, c, e,** Scatter plots depict the change of viral RNA copies on the log 10 scale over time. Each point represents a sample collected for an individual on a specific date. **b, d, f,** Boxplots present the comparison of viral RNA copies on the log 10 scale by Omicron subvariants. The Kruskal-Wallis p-value indicates the global comparison among the three subvariants. None of the pairwise comparison between two subvariants is significant at the level of 0.05. The *n* indicates the number of samples included in each boxplot. **a**, **b**, Fine EBA (≤5 µm in diameter); **c**, **d**, Coarse EBA (>5 µm in diameter); **e**, **f**, Total EBA (fine and coarse combined).

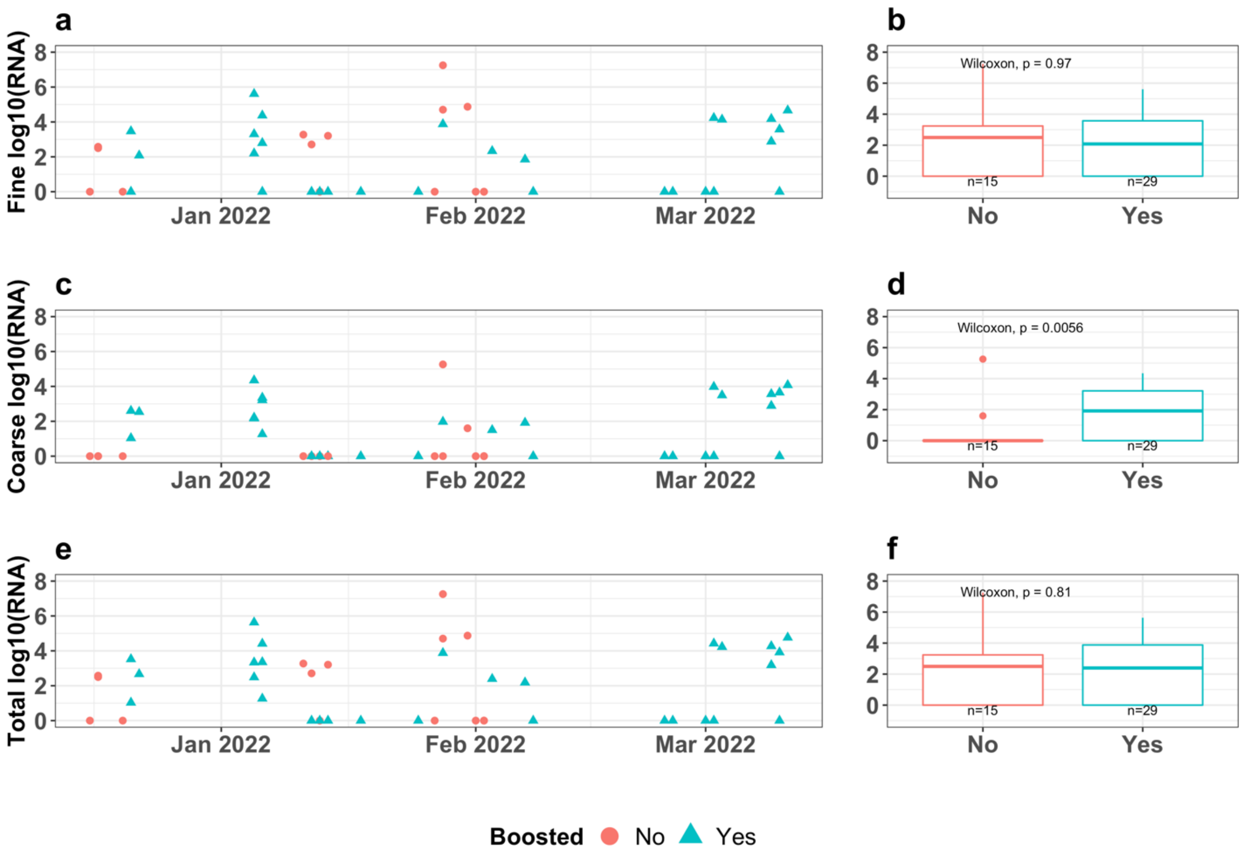


**Extended Data Figure 3. Viral RNA load in exhaled breath aerosol samples by booster status for Omicron cases, December 16, 2021 – March 11, 2022.**

**a, c, e,** Scatter plots depict the change of viral RNA copies on the log 10 scale over time. Each point represents a sample collected for an individual on a specific date. **b, d, f,** Boxplots present the comparison of viral RNA copies on the log 10 scale by booster status. The *n* indicates the number of samples included in each boxplot. **a**, **b**, Fine EBA (≤5 µm in diameter); **c**, **d**, Coarse EBA (>5 µm in diameter); **e**, **f**, Total EBA (fine and coarse combined).

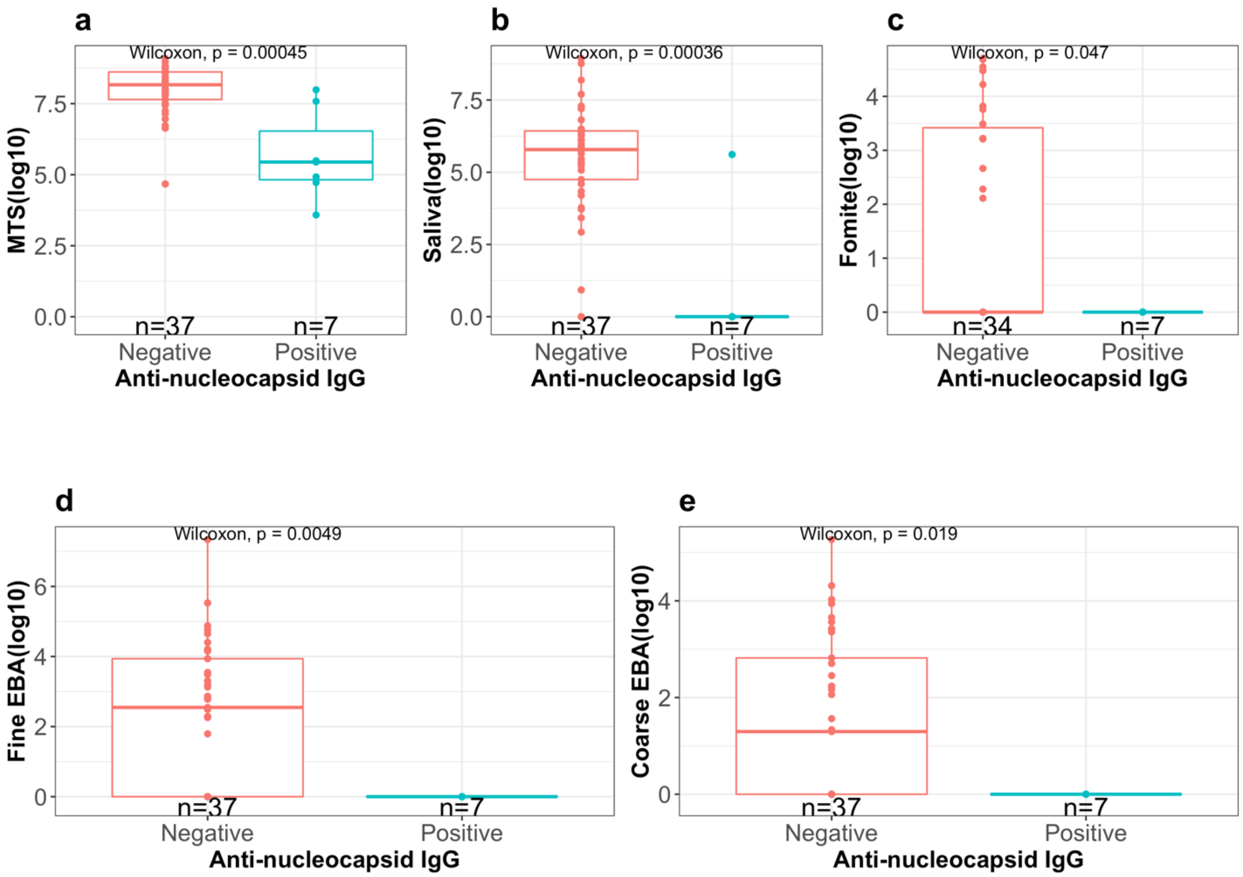


**Extended Data Figure 4. Viral RNA copies (log 10 scale) by status of anti-nucleocapsid IgG for Omicron cases, December 16, 2021 – March 11.**

Boxplots present the comparison of viral RNA copies on the log 10 scale by the status of anti-nucleocapsid IgG at baseline for 29 Omicron cases (24 negative, 5 positive). The *n* indicates the number of samples included in each boxplot. **a,** Mid-turbinate swab (MTS); **b,** Saliva; **c,** Fomite (swab of participant's mobile phone); **d** and **e,** *Fine* (≤5 µm in diameter) and *Coarse* (>5 µm in diameter) exhaled breath aerosol (EBA) from 30-minute sampling events. The *n* at the bottom of the plots indicates the number of samples.

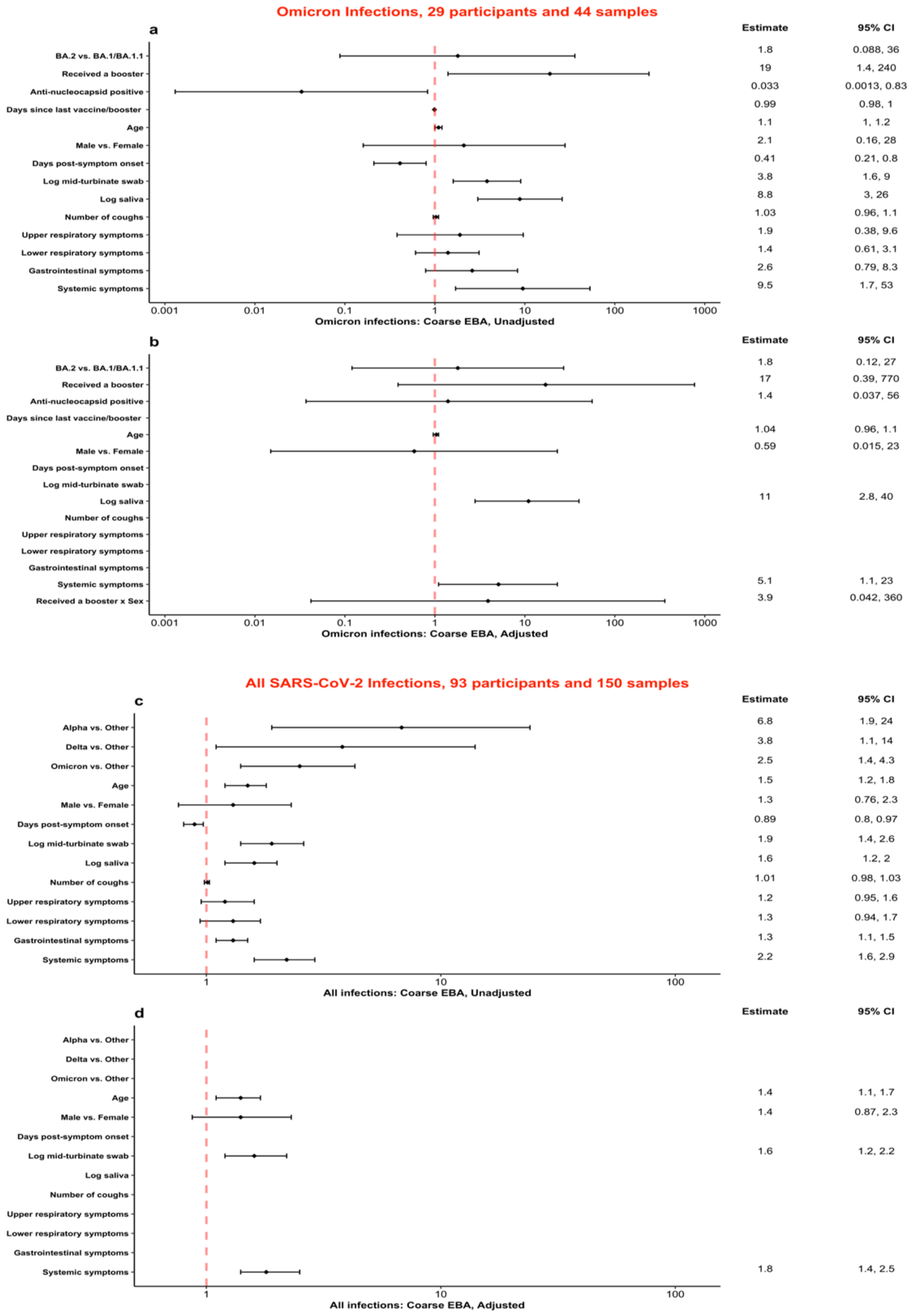


**Extended Data Figure 5. Predictors for SARS-CoV-2 RNA loads in *coarse* exhaled breath aerosol.**

**a-b, P**redictors for viral RNA loads in *coarse* exhaled breath aerosol among 29 participants with Omicron infections enrolled from December 16, 2021 to March 11, 2022. **c-d,** Predictors of viral RNA loads in *coarse* exhaled breath aerosol over the course of the pandemic from June 6, 2020 to March 11, 2022.

Effect estimates and their 95% confidence intervals from linear mixed effect models (observations censored by the limit of detection coded as 1) are shown as the ratio of RNA copy number of samples: variant to variants other than Alpha/Delta/Omicron, Omicron BA.2 to Omicron BA.1 and BA.1.1, received to not received a booster, anti-nucleocapsid positive to negative, male to female, or as the fold-increase in RNA copy number for a 10-year increase in age, 1-day increase in day post-symptom onset or days since last vaccine/booster, 1-count increase in numbers of coughs, and an interquartile range change in symptom scores, mid-turbinate swab and saliva RNA copy number.

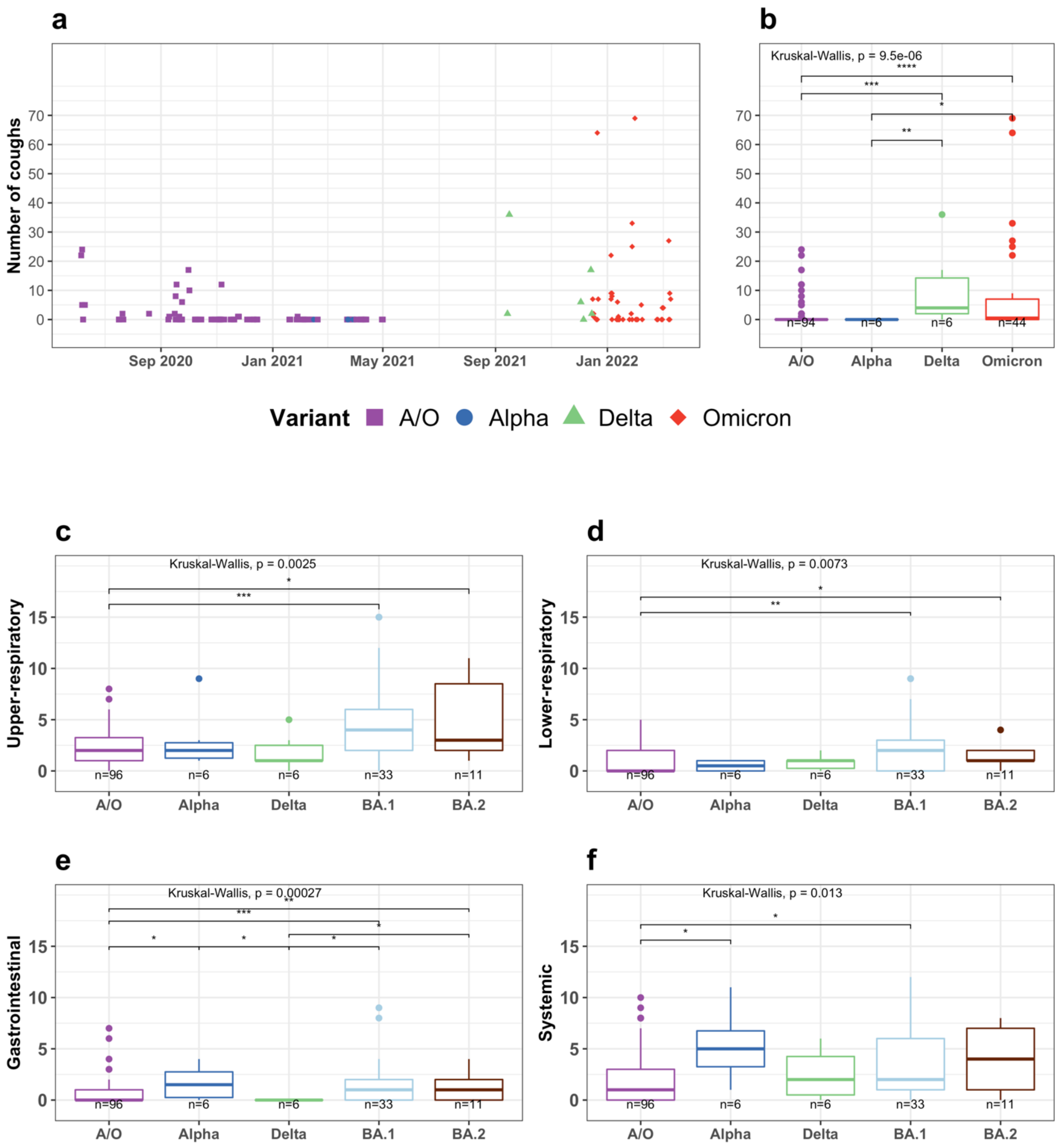


**Extended Data Figure 6. Number of coughs per 30-minute sampling and composite symptom scores for SARS-CoV-2 variants over time, June 6, 2020 – March 11, 2022.**

**a**, Each point in the scatter plot represents a sample collected for an individual on a specific date. **b,** The boxplots present the comparison of number of coughs per 30-minute sampling session by SARS-CoV-2 variants. **c - f,** The boxplots present the comparison of four composite symptom scores by SARS-CoV-2 variants/subvariants. The Kruskal-Wallis p-value indicates the global comparison among the four (**a-b**) or five (**c - f**) variants/subvariants. The asterisks indicate the pairwise comparison. Only those with a p-value less than 0.05 are shown (*: p <= 0.05; **: p <= 0.01; ***: p <= 0.001; ****: p <= 0.0001). The *n* indicates the number of samples included in each boxplot. *A/O* means SARS-CoV-2 ancestral strains and other variants not associated with increased transmissibility.

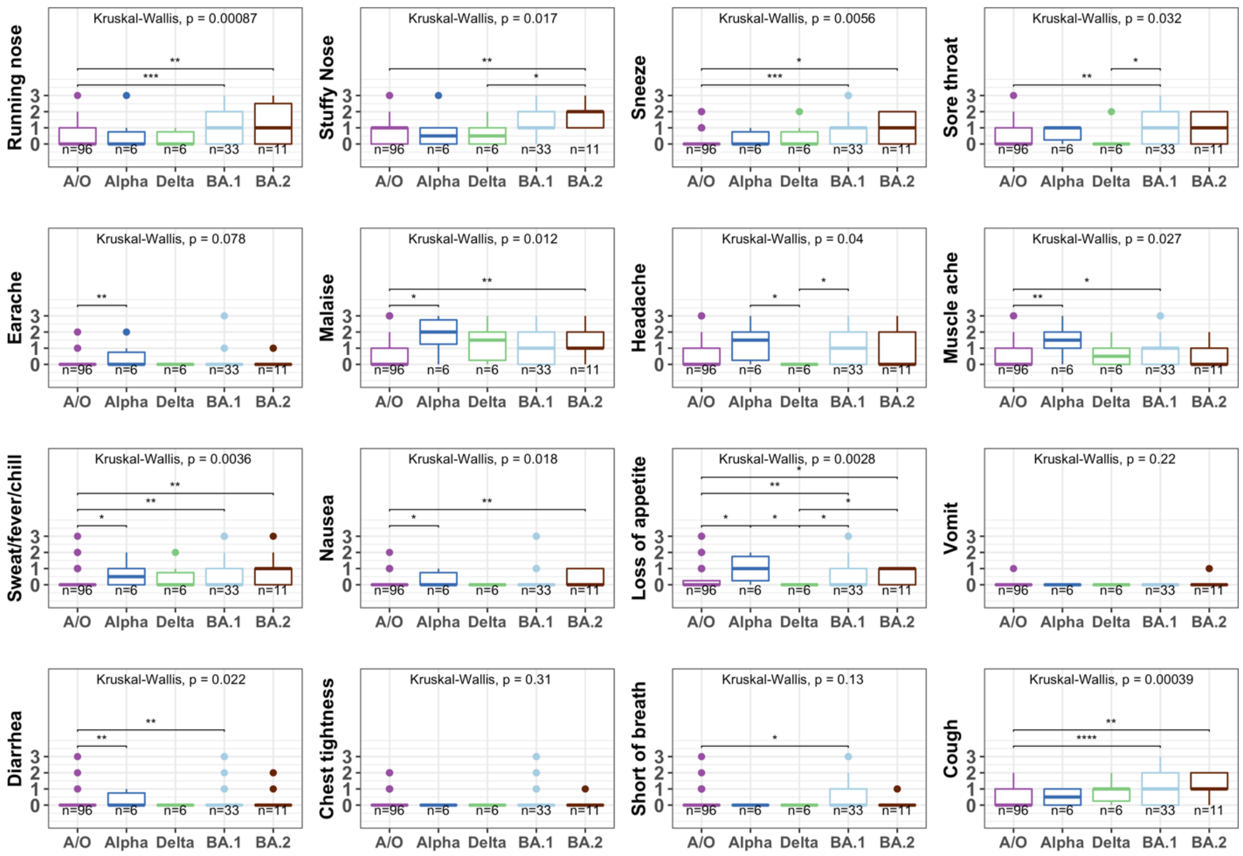


**Extended Data Figure 7. Self-reported symptoms for SARS-CoV-2 variants over time, June 6, 2020 – March 11, 2022.**

The boxplots present the comparison of 16 self-reported symptoms (on a scale of zero to three) by SARS-CoV-2 variants/subvariants. The Kruskal-Wallis p-value indicates the global comparison among the five variants/subvariants. The asterisks indicate the pairwise comparison. Only those with a p-value less than 0.05 are shown (*: p <= 0.05; **: p <= 0.01; ***: p <= 0.001; ****: p <= 0.0001). The *n* indicates the number of samples included in each boxplot. *A/O* means SARS-CoV-2 ancestral strains and other variants not associated with increased transmissibility.
